# Rare homozygous cilia gene variants identified in consanguineous congenital heart disease patients

**DOI:** 10.1101/2023.08.25.23294614

**Authors:** Daniel A. Baird, Hira Mubeen, Canan Doganli, Jasmijn B. Miltenburg, Oskar Kaaber Thomsen, Zafar Ali, Tahir Naveed, Asif ur Rehman, Shahid Mahmood Baig, Søren Tvorup Christensen, Muhammad Farooq, Lars Allan Larsen

## Abstract

**BACKGROUND:** Congenital heart defects (CHD) appear in almost one percent of live births. Asian countries have the highest birth prevalence of CHD in the world. Recessive genotypes may represent a significant CHD risk factor in Asian populations, because Asian populations have a high degree of consanguineous marriages, which increases the risk of CHD. Genetic analysis of consanguineous families may represent a relatively unexplored source for investigating CHD etiology.

**METHODS:** To obtain insight into the contribution of recessive genotypes in CHD we analysed a cohort of forty-nine Pakistani CHD probands, originating from consanguineous unions. The majority (82%) of patient’s malformations were septal defects. We identified protein altering, rare homozygous variants (RHVs) in the patient’s coding genome by whole exome sequencing.

**RESULTS:** The patients had a median of seven damaging RHVs each, and our analysis revealed a total of 758 RHVs in 693 different genes. By prioritizing these genes based on variant severity, loss-of-function intolerance and specific expression in the developing heart, we identified a set of 23 candidate disease genes. These candidate genes were significantly enriched for genes known to cause heart defects in recessive mouse models (P<2.4e-06). In addition, we found a significant enrichment of cilia genes in both the initial set of 693 genes (P<5.4e-04) and the 23 candidate disease genes (P<5.2e-04). Functional investigation of *ADCY6* in cell- and zebrafish-models verified its role in heart development.

**CONCLUSIONS:** Our results confirm a significant role for cilia genes in recessive forms of CHD and suggest important functions of cilia genes in cardiac septation.

## INTRODUCTION

Congenital heart defects (CHD) comprise simple to complex heart malformations, which affect up to one percent of live births ^1, 2^. Approximately 90% of CHD patients survive to adulthood and an estimated 12 million people worldwide are currently living with CHD ^3, 4^.

A recent meta-analysis of the global birth prevalence of CHD suggested that the prevalence of CHD is significantly higher in Asia compared to the rest of the world ^1^. The etiology behind this observation is most likely complex, but many Asian countries have a high degree of consanguineous marriages ^5^ and several studies have shown that the risk of CHD is increased in children of consanguineous couples ^6^. Thus, it is possible that recessive genotypes (RGs) segregating in the population may represent a significant CHD risk factor in Asian countries with a high degree of consanguineous unions.

Several autosomal recessive monogenic syndromes are known, where CHD represent a part of the clinical spectrum ^7^. However, only a limited number of studies have been conducted to identify variants, genes and molecular mechanisms associated with autosomal recessive inherited CHD in the general population. Analysis of whole-exome sequencing (WES) data from 2,645 parent–offspring trios suggested that RGs may account for nearly 2% of CHD ^8^. Furthermore, a subsequent study based on the same cohort, showed that genes involved in cilia structure or function (cilia genes) are significantly enriched for RGs in CHD probands, particularly in patients with heterotaxy/laterality defects ^9^. Cilia are thin microtubule-based antenna-like organelles that project from the surface of most cells ^10^. In the developing embryo, cilia play major roles in establishing left-right asymmetry and detecting and conveying cellular signalling, which control organ and tissue development, including the heart ^11, 12^. The association between RGs and cilia genes in CHD is corroborated by WES studies of CHD patients with heterotaxy/laterality defects or other severe heart malformations ^13, 14^ and from the result of a recessive forward genetic screening of mice ^15^.

There is a paucity of genetic studies on autosomal recessive CHD patients, with only a few studies conducted ^13, 16^. Therefore, genetic analysis of CHD patients from populations with high frequency of consanguinity represent a relatively unexplored source for identification of novel CHD disease genes and mechanisms. Pakistan has one of the highest rates of consanguineous marriages in the world, with over 50% of marriages between second cousins or closer relatives ^5^. In the current project, we used WES to analyze a cohort of Pakistani CHD probands originating from consanguineous unions.

## METHODS

Full methods are available in the Supplemental Material. The data that support the findings of this study are available from the corresponding authors upon reasonable request. Individual exome sequencing data cannot be shared due to concerns over patient privacy. The study was approved by the Institutional Review Board GC University Faisalabad, Faisalabad, Pakistan and the Ethical Review Committee (ERC), Punjab Institute of Cardiology, Lahore, Pakistan. Informed consent was obtained from all participating individuals or their parents for the collection of blood samples, genetic analyses, and publication of genetic information.

## RESULTS

We analysed 49 CHD patients (20 females, 29 males) from 48 consanguineous Pakistani families. Parents of the patients did not have CHD. Age of the patients ranged from 1 year to 53 years. None of the patients had previously been diagnosed with a genetic disorder. The CHD diagnoses covered VSD (57.1%), ASD (24.5%), TOF (8.2%) and others (10%) (Table 1).

**Table 1.**
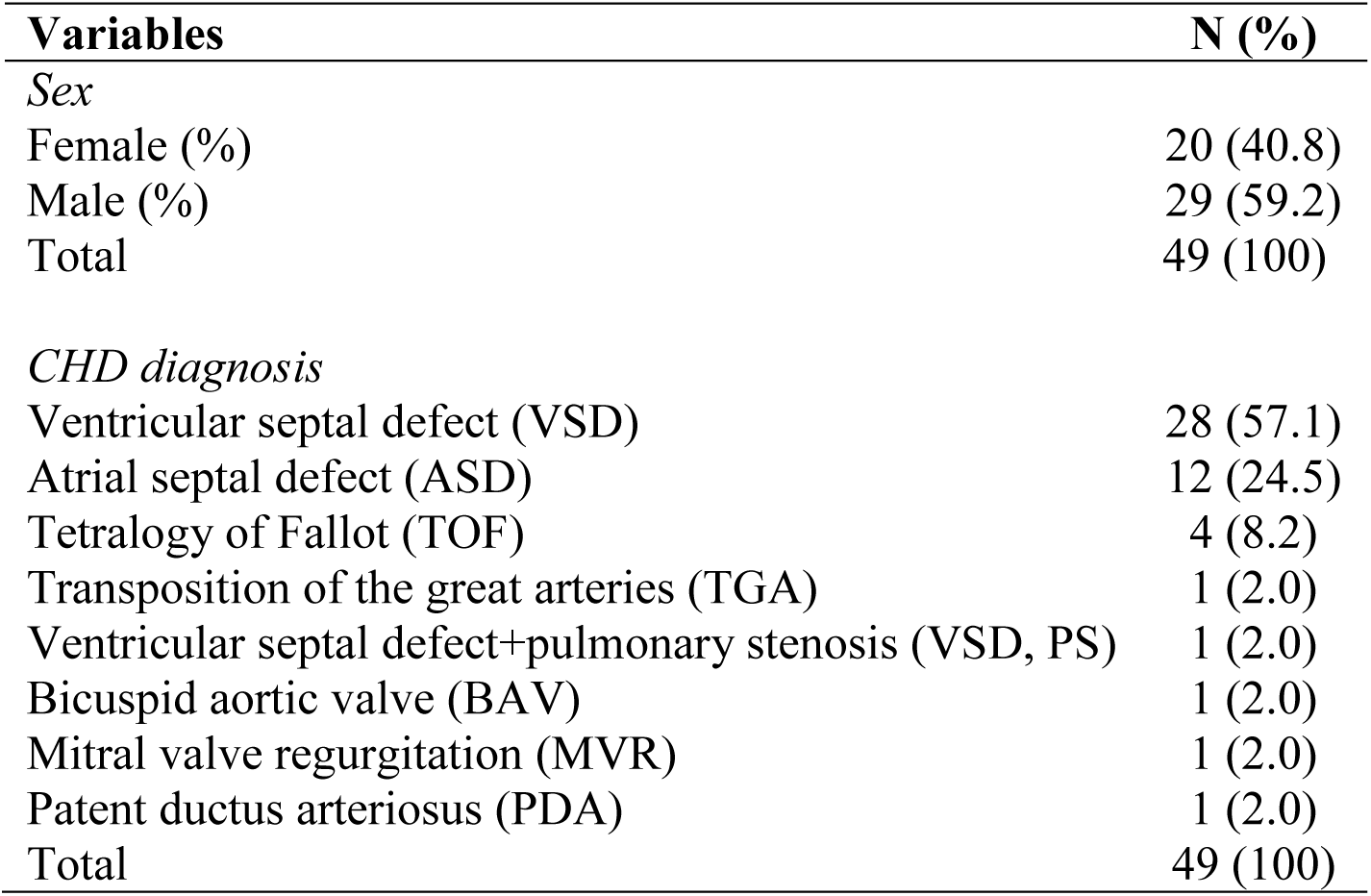
Clinical characteristics of patients.

We identified variants in the coding sequence of the patient’s genome by whole exome sequencing (WES). Analysis of WES data revealed large regions of homozygosity-by-descent (HBD) in the patient’s genome (Fig. 1A). The genome-wide size of HBD ranged from 5.1 to 469.7 Mbp per sample, with a median value of 167.9 Mbp (5.6% of the genome) (Fig. 1B). We identified a total of 1,168 HBD regions, each encompassing at least 50 homozygous variants. The individual HBD regions ranged from 66.1 Kbp to 71.2 Mbp in size, with a median value of 4.96 Mbp (Fig. 1C). The longest HBD segment per sample was 30.2 ± 14.8.

**Figure 1.**
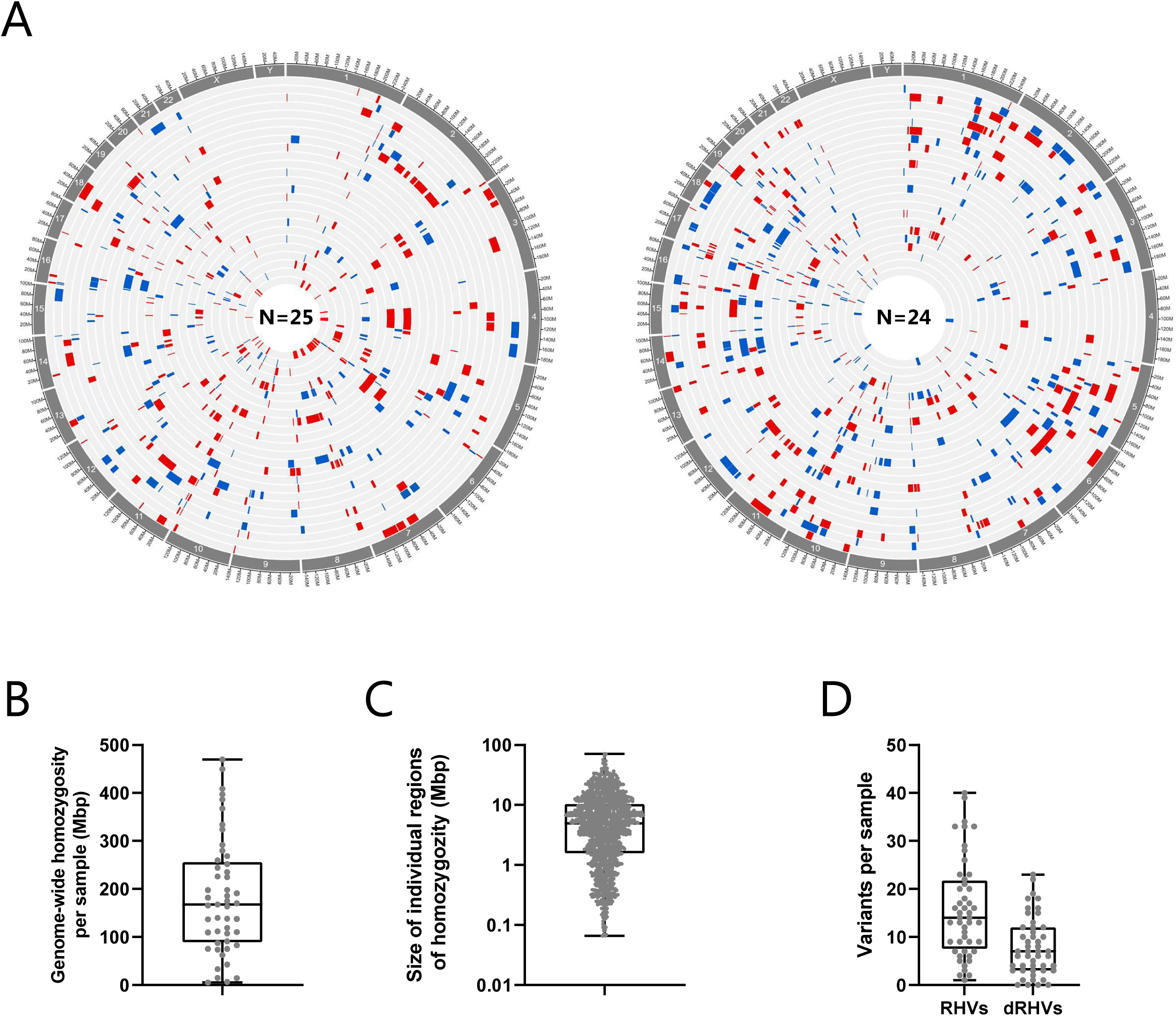
Distribution of rare homozygous variants in CHD patients from consanguineous families. **A.** Genomic regions of homozygosity (ROH) in 49 patients. The genomic localization of ROH, containing at least 50 homozygous variants, is shown with red or blue bars. Each circle represents the genome of one patient. The size of each ROH is shown in (C). **B.** Total size of ROH per patient in mega base-pairs (Mbp). **C.** The size distribution of all 1,167 ROH identified in patients. **D.** The number of rare homozygous variants (RHVs) and damaging RHVs (dRHVs) identified per sample.

By filtering of the WES data, we identified a total of 758 protein altering rare homozygous variants (RHVs) in 693 candidate disease genes (gene-set 1, GS1) (Table S1). Thirty-one (4.1%) of the variants were protein truncating (nonsense or frameshift variants) and 695 (91.7%) of the variants were missense variants. The number of RHVs per sample ranged from one to 40, with a median value of 14 and while the number of damaging RHVs (CADD score ≥21) per sample ranged from zero to 23, with a median value of 7 (Fig. 1D).

We hypothesized that if RHVs are causative of CHD in our patient cohort, we would expect that GS1 was enriched for known CHD genes. To test this hypothesis, we calculated the overlap between the 693 genes in GS1 and curated lists of genes known to cause CHD in mouse models and patients (Table S2) ^17, 18^. We observed significant enrichment of human genes causing biallellic (recessive) CHD and genes from mouse models of CHD (most of which are recessive models), but we observed no enrichment of human genes causing monoallellic (dominant) CHD (Figure S1). The enrichment of genes known to cause recessive CHD in GS1 support that rare RHVs are associated with CHD in our patient cohort. A high CADD score of a given variant indicates that the variant is more likely to be deleterious to the gene product ^19^, while a low LOEUF score indicates that a gene is less tolerant to loss-of-function ^20^. Thus, as a measure of severity of the RHVs and the intolerance towards loss-of-function, we determined the CADD score of each variant and the LOEUF score of each gene in GS1. The distribution of CADD scores and LOEUF scores of RHVs and GS1 genes, respectively are shown in Fig. 2A. We used the median value of the CADD scores (21) and LOEUF scores (0.83), as a simple cut-off for variant severity and loss-of-function intolerance, and thus define variants with CADD score ≥ 21 as likely damaging and genes with LOEUF ≤ 0.83 as likely intolerant to loss-of-function. Next, we grouped GS1 into three subsets: 185 genes with low likelihood of causing CHD (GS1a, CADD <21, LOEUF>0.83), 323 genes with medium likelihood of causing CHD (GS1b, CADD≥21, LOEUF>0.83 and CADD<21, LOEUF≤0.83) and 192 genes with high likelihood of causing CHD (GS1c, CADD≥21, LOEUF≤0.83). The CADD score and LOEUF of the three gene subsets are plotted in Fig. 2B and listed in Table S1.

**Figure 2.**
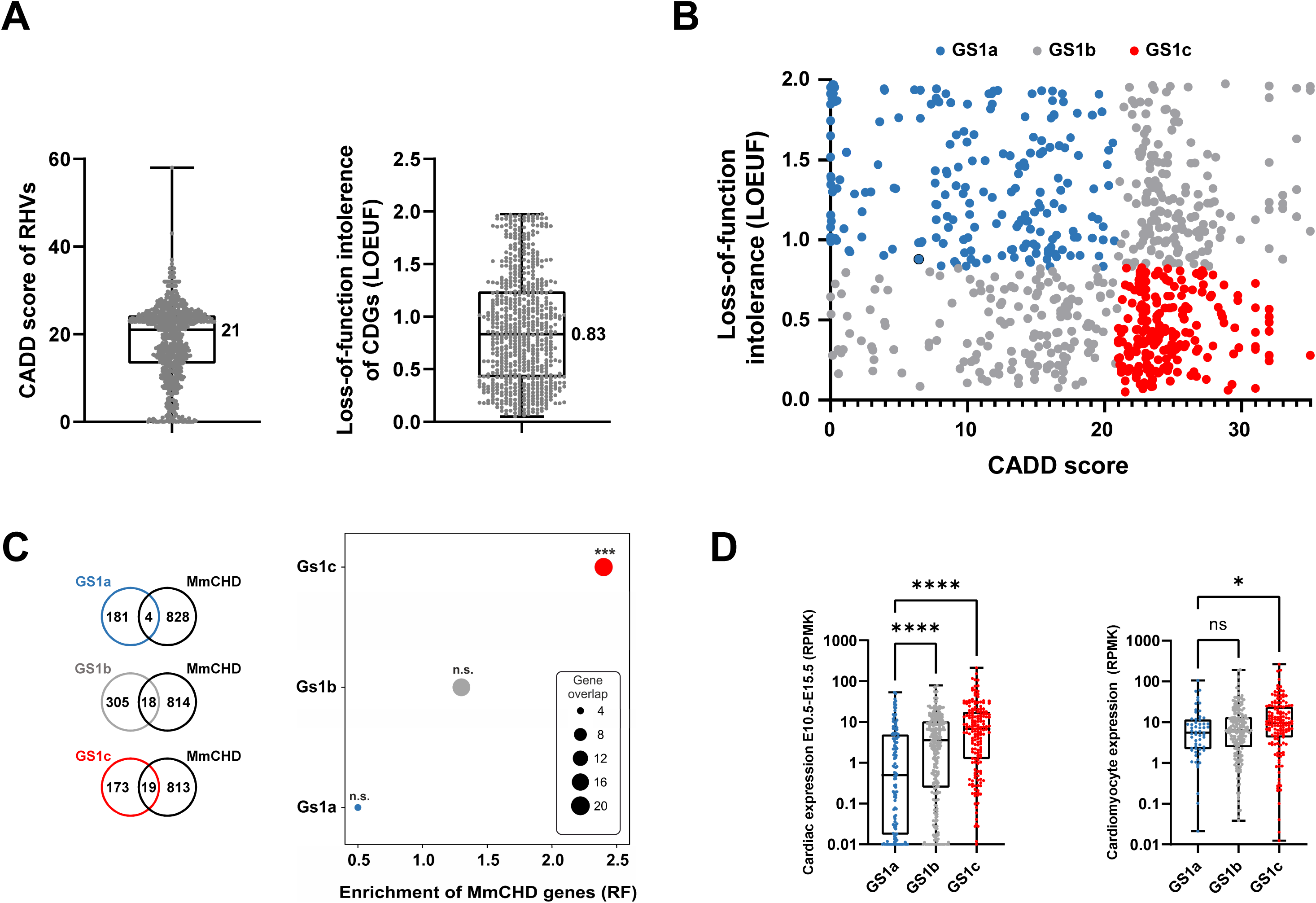
Prioritization of candidate disease genes. **A**. Distribution of CADD scores of RHVs identified in the patients (left) and distribution of loss-of-function intollerence score (LOEUF) in candidate disease genes (CDGs, right). Median values are indicated. **B**. X-Y plot of 678 CDGs containg rare homozygous variants (geneset 1, GS1). The genes are plottet according to CADD score of RHVs identified in each gene (X-axis) and loss-of-function intolerence of the gene (LOEUF, Y-axis). GS1 was divided in three sub-groups. Low-likelihood CDGs (GS1a, blue): CADD<21, LOEUF>0.83. Medium likelihood CDGs (GS1b, grey): CADD<21 and LOEUF≤0.83 or CADD≥21 and LOEUF>0.83. High-likelihood CDGs (GS1c, red): CADD≥21 and LOEUF≤0.83. **C**. Enrichment of MmCHD genes among three CDG subgroups. Enrichment was calculated by comparing gene-overlap between MmCHD genes and the three sub-groups of GS1. Enrichment is shown as representation factor (RF). A hypergeometric distribution was used to test the significance of the overlaps. **D**. Expression level of the three GS1 subsets in mouse embryonic hearts (left) ^22^ and in vitro cultures of cardiomyocytes (right) ^21^. Difference between medians were determined using ANOVA (Kruskal-Wallis test). Asterisks indicate P-values: * P<0.05, *** P<0.001, **** P<0.0001. ns: not significant.

To validate the categorization of GS1 into three gene-sets based on their likelihood of causing CHD, we calculated the enrichment of CHD genes known from recessive mouse models (Table S2) within each GS1 subset and observed that only the GS1c subset is significantly enriched for such genes (RF=2.4, P= 4.3e-04, Fig. 2C). In addition, we performed the same enrichment analysis using a human gene-set obtained from clinical exome sequencing (CES) of CHD patients from a consanguineous cohort (Table S2) ^16^. Analysis of the much smaller human gene-set gave similar results, with enrichment for the GS1c subset only (RF=8.3, P=0.001, Figure S2). Finally, we utilized publicly available transcriptomic datasets ^21, 22^ to compare the transcription level of genes in mouse E10.5-E15.5 embryonic hearts and cardiomyocytes differentiated from mESC, in each GS1 gene-set (Fig. 2D). For both data materials, the median expression value of GS1c was significantly higher than the median value of GS1a, while the median value of GS1b displayed an intermediate level of expression, compared to the other groups. Based on these analyses, we conclude that GS1c is enriched for CHD disease genes, compared to the other two gene-sets.

We performed a final prioritization of the 192 genes in GS1c by examining their specific expression in the developing heart. To this end, we used RNAseq data from mice (E10.5-E18.5) to compare gene expression in the heart with gene expression in brain (Br), liver (Li) and kidney (Ki). The analysis identified 23 genes with at least two-fold higher expression in the heart compared to the other three tissues during development (Table 2, Figure S3). These 23 genes are enriched for MmCHD genes (RF=8.4, P < 2.4e-06) and represent a list of high-likelihood candidate disease genes (CDGs) for CHD in our probands.

**Table 2.**
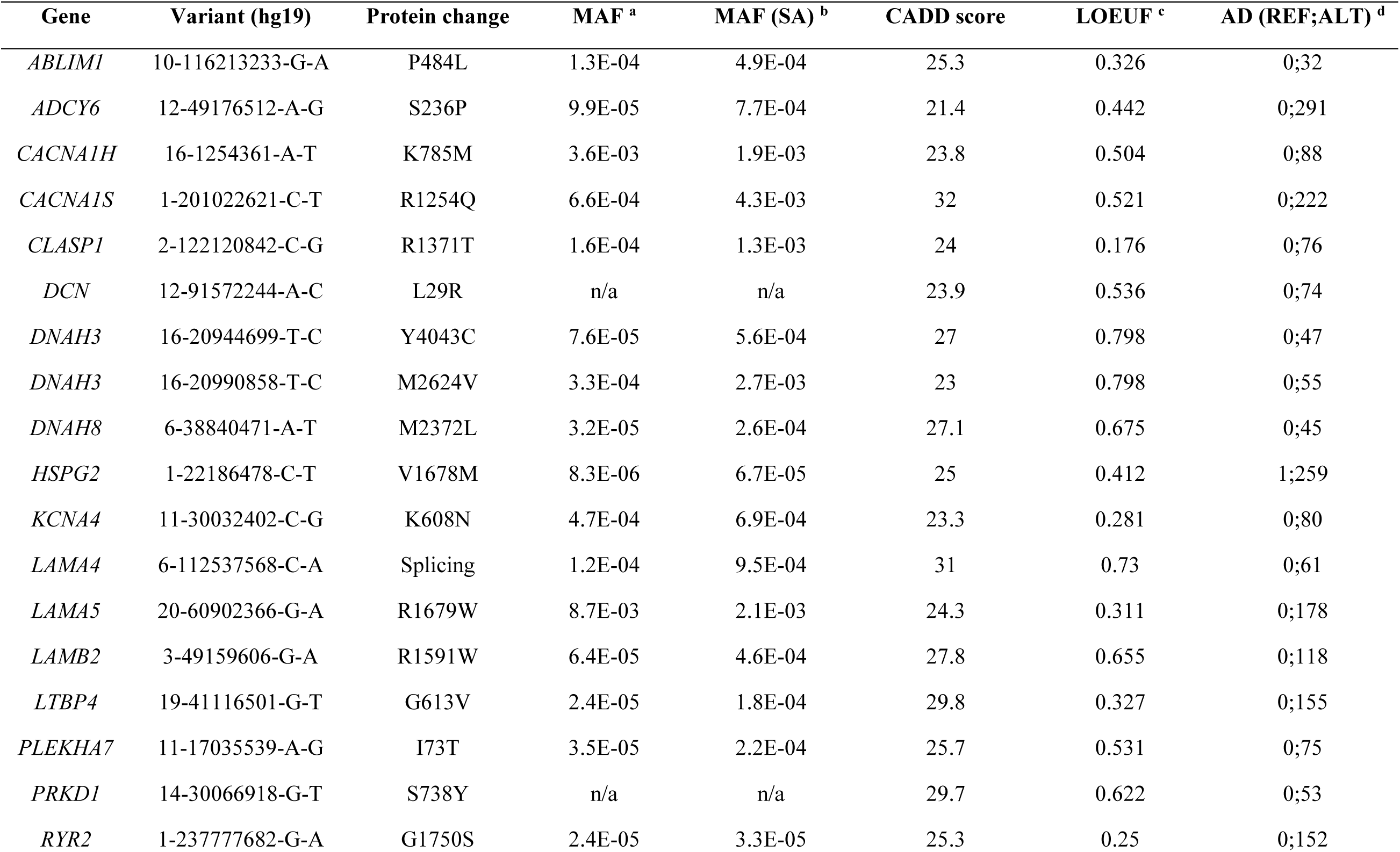
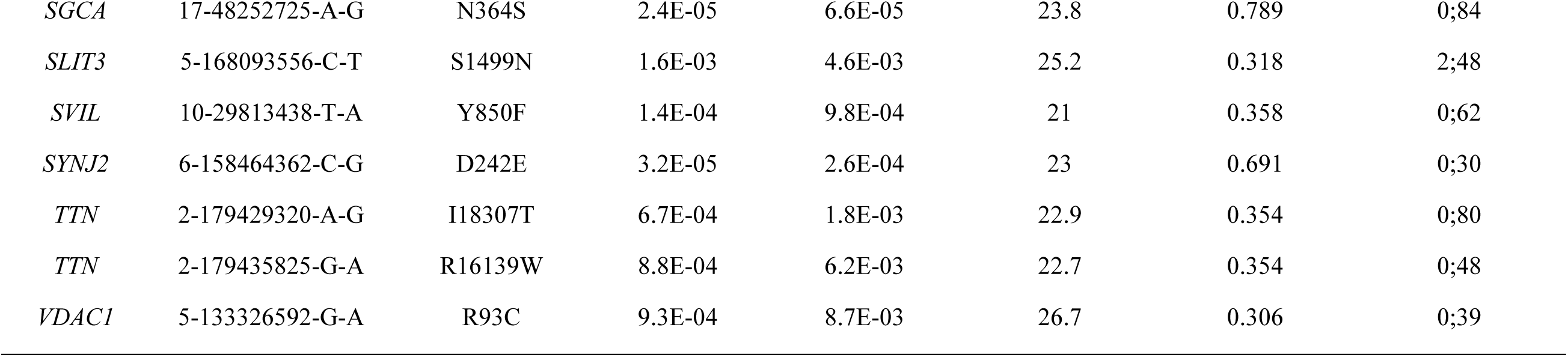
Rare homozygous variants identified in 23 candidate disease genes. RHVs in all 693 genes in GS1 are shown in supplementary table S2. ^a^ Minor allele frequency (MAF) among all 125,748 exomes in GnomAD. ^b^ MAF among 15,308 South Asian exomes in GnomAD. ^c^ Loss-of-function observed/expected upper bound fraction (LOEUF). ^d^ Allelic depth (AD, number of sequencing reads covering the variant) of variant in reference genome (REF) and alternative variant (ALT).

Previous analysis of data obtained from exome sequencing of parent-patient trios revealed that cilia genes are enriched for damaging rare recessive genotypes, suggesting that cilia genes may serve as a reservoir of rare recessive variants that can cause CHD in homozygous or compound heterozygous form ^9^. This observation prompted us to investigate our gene-sets for ciliary gene enrichment. We calculated the overlap between CiliaCarta, a compendium of 935 unique cilia genes, and the 693 genes in GS1. We observed significant enrichment of cilia genes in GS1 (RF=1.6, P<5.4e-04, 52 cilia genes).

We proceeded to compare the frequency of individuals with RHVs in each of the 52 cilia candidate genes between our patient cohort and controls from the GnomAD database (Fig. 3A). This comparison showed a ten-fold higher frequency of individuals with RHVs in cilia genes, among our patients, compared to the control populations. Additionally, we compared the CADD scores of RHVs in the 52 cilia genes, with the 641 non-cilia genes in GS1 and observed significantly higher median CADD score for variants in the cilia genes (Fig. 3B). Furthermore, we observed enrichment of damaging variants (CADD score ≥21) in the cilia genes (Fig. 3C).

**Figure 3.**
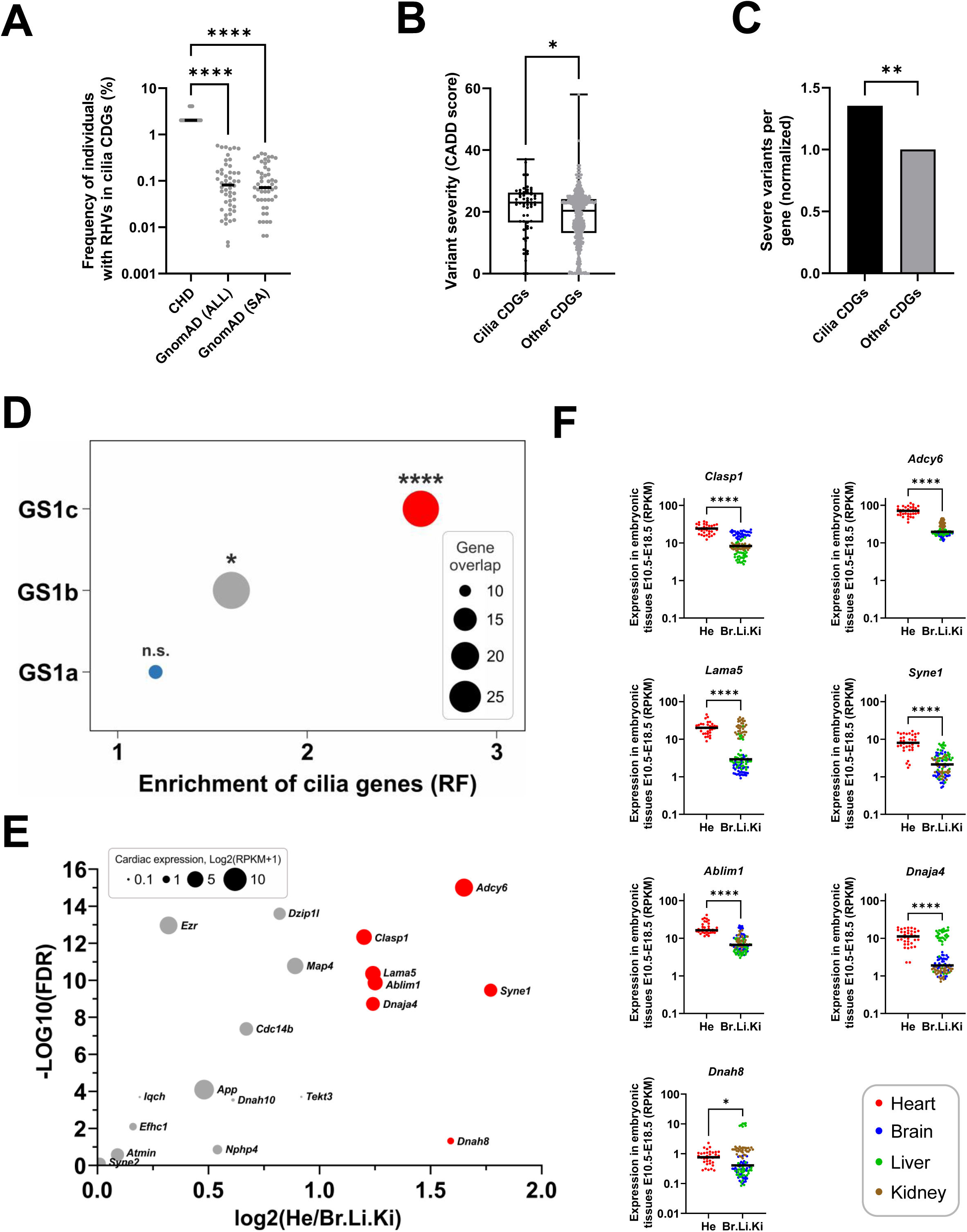
Cilia genes are enriched for rare homozygous variants. **A**. Frequency of individuals with RHVs in 52 cilia CDGs. Frequency per gene in patient cohort (N=49) was compared with frequency in GnomAD populations. GnomAD (All): Total GnomAD sample (N=125,748). GnomAD (SA): South Asian GnomAD sub-sample (N=15,308). Statistic comparison was performed using ANOVA (Kruskal-Wallis test). **B**. Comparison of variant severity of RHVs identified in cilia CDGs (N=52) and other CDGs (N=641). Variant severity is indicated as CADD score. Difference between medians were determined using ANOVA (Kruskal-Wallis test). **C**. Normalized comparison of severe RHVs per gene, between cilia CDGs and other CDGs. Severe variants were defined as RHVs with CADD score ≥ 21. Statistical significance of the difference was determined using Fisher’s exact test. **D**. Enrichment of cilia genes among three subsets of GS1. Enrichment was calculated by comparing gene-overlap between CiliaCarta genes (N=935) and genes within each GS1 subset. Enrichment is shown as representation factor (RF). Hypergeometric statistics was used to test the significance of the overlaps. **E**. Vulcano plot of 52 cilia genes showing the differene in gene expression between developing heart (He) and developing brain (Br), Liver (Li) and Kidney (Ki) in mice at E10.5-E18.5 ^22^. X axis shows the log2 difference between average expression in He and average expression in Br, Li and Ki (only positive values shown). Y axis shows the significance, calculated as –Log10 to the false discovery rate (FDR) (Mann-Whitney U test, adjusted for multiple testing). Significant genes, with log2 difference >1 is shown with red color. The size of the circle indicate log2 of the average expression of the gene in developing hearts. **F**. Tissue comparative expression of individual genes with fold change >1. Asterisks indicate P-values: * P<0.05, ** P<0.01, **** P<0.0001. ns: not significant.

We hypothesized that if RHVs in cilia genes are causative in our patient cohort, then we would expect the largest enrichment of cilia genes in the GS1c gene-subset, compared to the other two gene-subsets. To test this, we calculated the enrichment of cilia genes in all three GS1 gene-subsets. We did not observe significant enrichment in the GS1a gene-subset, but both GS1b and GS1c were enriched for cilia genes, with RF of 1.6 and 2.6, respectively (Fig. 3D) and we observed the strongest enrichment for cilia genes in the final subset of 23 CDGs (RF=5.6, p < 5.2e-04). When we compared the frequency of RHVs in each of 23 cilia genes in GS1c, we observed a very high frequency in our patients compared to GnomAD controls (Figure S4). Overall, these analyses support our hypothesis that RHVs in cilia genes are associated with CHD in our patient cohort.

To identify cilia candidate genes with heart-specific expression, we again used publicly available RNAseq data from mice (E10.5-E18.5) ^22^ and compared gene expression in the heart with gene expression in brain, liver and kidney for all 52 cilia genes within GS1 (Fig. 3E, F). We identified seven cilia genes of which the mouse orthologue had more than two-fold higher expression in the developing heart compared to brain, liver and kidney. Of these seven cilia genes, five are within the list of 23 CDGs (Table 2); *ABLIM1, ADCY6, CLASP1, DNAH8, LAMA5*.

Recently, we have identified *ADCY2* and *ADCY5* as novel CHD disease genes ^17^, thus *ADCY6* appeared to be an interesting disease candidate. In addition, *Adcy6* exhibited the most significant difference in expression between heart and other tissues and the highest expression in the embryonic heart among the ciliary candidate disease genes (Fig 3E, F). However, *ADCY6* has not previously been associated with CHD or heart development. Therefore, we used cell models and zebrafish to investigate a cilia-related function of ADCY6 in heart development.

We and others have previously shown that P19CL6 cells is a simple and valid model to study early cardiomyogenesis ^23–26^. P19CL6 are teratocarcinoma derived pluripotent stem cells, which will spontaneously differentiate into beating cardiomyocytes within 12-14 days after addition of DMSO to the growth media. During the differentiation of P19CL6 cells, a decreased expression of the stem cell marker SOX2 is concomitant with increased expression of the cardiomyocyte transcription factor GATA4 (Figure S5A). Around day 12, development of sarcomeric structures can be evidenced by a striated pattern of Troponin I (alongside α-actinin) and spontaneously beating cardiomyocytes start to appear in the culture (Figure S5B).

To investigate whether ADCY6 has a potential ciliary function during cardiomyogenesis, we utilized immunofluorescence microscopy to examine ciliary localization of ADCY6 at four time points during differentiation of P19CL6 cells into cardiomyocytes. At day 0, when DMSO is added, ADCY6 was mainly localized to the base of the primary cilium in the stem cells (Figure 4A,B). However, at two later stages during the differentiation process (day 7, 10), we observed a gradual two-fold increase in the levels of ADCY6 protein at the primary cilium, with ADCY6 localized along the length of the cilium (Figure 4A-C). At day 12, the amount of ADCY6 was decreased to a level similar to day 7. These data demonstrate that ADCY6 localize to the primary cilium in a temporal manner during cardiomyogenesis.

**Figure 4.**
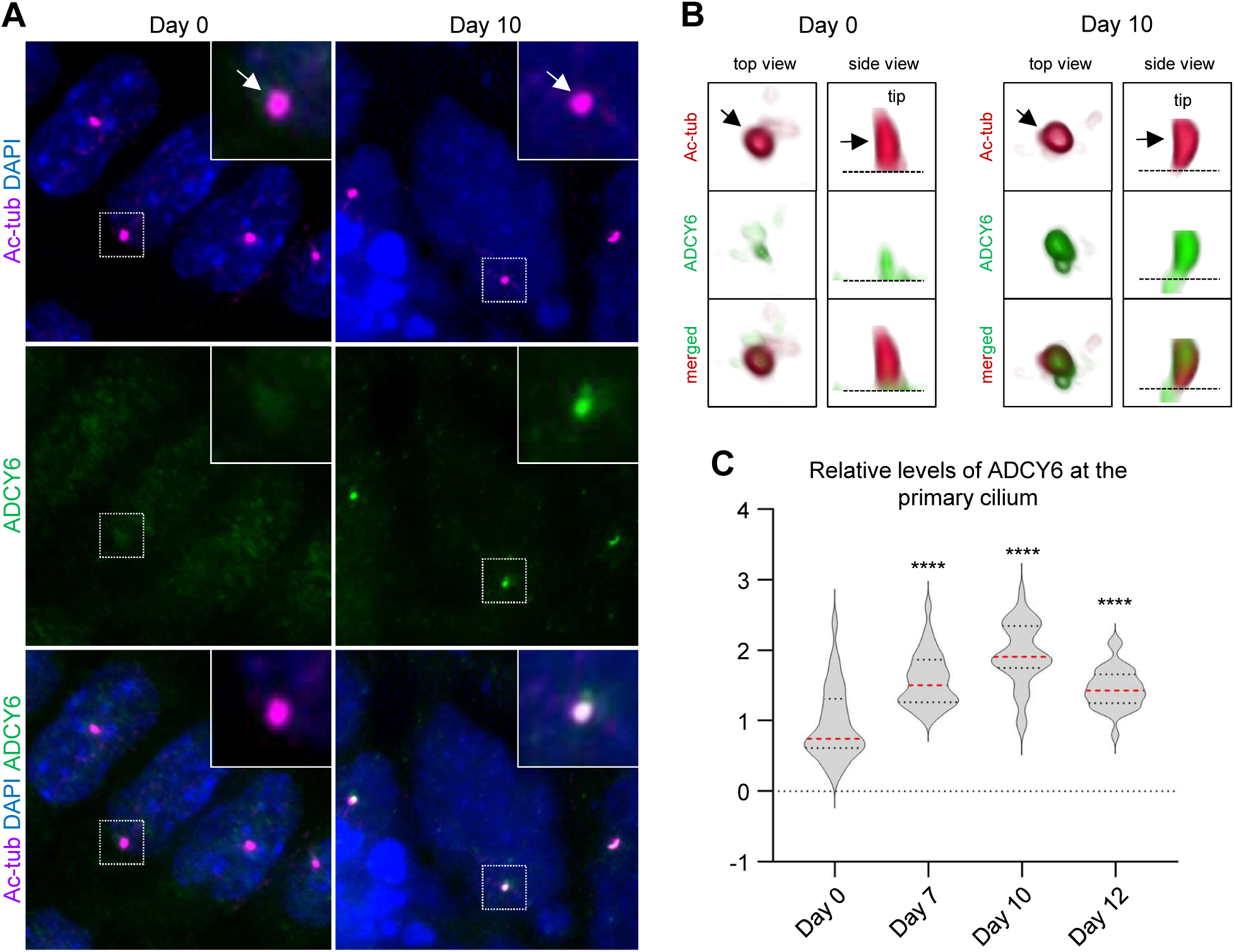
Temporal localization of ADCY6 to primary cilia during cardiomyogenesis. ADCY6 accumulate at primary cilia during differentiation of P19.CL6 cells into cardiomyocytes. **A**. Representative images P19.CL6 cells at day 0 (stem cells) and day 10 (cardiomyocytes). Arrows point to primary cilia. Scale bar, 10 µm. **B**. 3D visualization of representative cilia at day 0 and day 10 of differentiation. Arrow points to the primary cilium. Asterisk marks the ciliary base. **C.** Violin plots of quantification of ADCY6 fluorescence levels at primary cilia during days 0, 7, 10 and 12 of differentiation. Asterisks indicate P-values: ****P<0.0001.

To further study the function of *ADCY6* in heart development *in vivo*, we generated F0 mutant (crispant) models of the *ADCY6* zebrafish orthologues *adcy6a* and *adcy6b* by simultaneous targeting of four exons in each gene (Figure S6A). Genotyping with PCR primers showed that all guides produced efficient mutagenesis individually, indicating that simultaneous injection of all four guides leads to efficient gene knock-down (Figure S6B)^27, 28^. Crispants for *adcy6a* and *adcy6b* displayed cardiac edema, which was not observed in control samples, at 2 dpf (Fig. 4A), suggesting cardiac defects in both *adcy6* crispants.

To further delineate the cardiac phenotype of *adcy6a* and *adcy6b* crispants, we performed *in situ* hybridization (ISH) using a cardiac-specific *myl7* probe to label the myocardium and examined heart morphology. At 2 dpf, significant proportions of both *adcy6a* and *adcy6b* crispants displayed cardiac defects, compared to uninjected and *scramble* controls (Fig. 4B-D). A total of 39% and 63% of *adcy6a* and *adcy6b* crispants, respectively, presented with abnormal cardiac morphology as either reversed heart, straight heart, or other defects (Fig. 4B, E, F). In both *adcy6* orthologue crispants, a straight heart was the predominant phenotype. Only a small percentage of uninjected and *scramble* embryos displayed cardiac defects. These findings indicate that frameshift mutations in *adcy6a* and *adcy6b* independently disrupt cardiac morphology. To assess cardiac functionality, we measured the heart rate of crispant embryos at 2 dpf. Both *adcy6a* and *adcy6b* crispants, independently, presented with significantly lower heart rates when compared to control samples (Figs. 4G,H).

Finally, we co-injected guides targeting both *adcy6a* and *adcy6b* simultaneously (Figure S7). These results supported the involvement of *ADCY6* in heart development, but the frequency of heart defects did not exceed what we observed for *adcy6b* alone.

In summary, CRISPR-cas9 mediated functional analysis of *ADCY6* orthologues in zebrafish models support that ADCY6 have important functions in heart development.

## DISCUSSION

We studied a cohort of Pakistani CHD patients originating from consanguineous unions. WES analysis revealed that, on average, HBD regions covered more than 5% of the patient’s genome, thus confirming the expected consanguinity. The longest HBD segment per sample was 30.2 ± 14.8 and the median number of damaging RHVs per sample was 7. These values are comparable to reported in a recent WES analysis of Turkish CHD patients from consanguineous families ^13^. The majority of heart malformations in our patient cohort, however, were simple (septal defects) as compared to a majority of complex defects, including a large number of rare laterality defects, identified in the Turkish cohort ^13^.

Our approach for filtering variants was centered around the idea that a substantial portion of the genetic influence on CHD in our patient population is attributable to homozygosity for recessive variants that are deleterious. To this end, we filtered for RHVs and used a combination of variant classification and loss-of-function intolerance to identify a set of 192 genes (GS1c) with high likelihood of causing CHD in our patients. Enrichment analysis using datasets of known CHD disease genes confirmed that this strategy was meaningful. To further prioritize the candidate genes, we took advantage of a comprehensive transcription profiling dataset obtained from mouse embryos at different developmental stages. By comparing the cardiac transcription profile with the profile in brain, liver and kidney for each gene in GS1c, we identified 23 genes with specific expression in developing hearts. Of these 23 genes, eight are known to cause heart defects in mouse models (*CACNA1H*, *HSPG2*, *LAMA4*, *LAMA5*, *LTBP4*, *RYR2*, *SLIT3*, *TTN*) ^29^ while *PRKD1*, among the remaining 14 candidate genes, has previously been implicated in CHD. Two independent reports have identified rare homozygous loss-of-function *PRKD1* variants as causative in consanguineous families with multiplex CHD cases ^30, 31^. In addition, one case with complex CHD was found homozygous for a rare nonsense variant in *PRKD1* in a CES screen of 2,219 families from a highly consanguineous population ^16^. Interestingly, WES analysis of a large cohort of CHD patients, implicated rare *de novo* missense mutation in *PRKD1* in CHD, thus suggesting that *PRKD1* variants can have both recessive and dominant effects on CHD ^32^. Our patient was homozygous for a S738Y missense variant within the protein kinase domain of *PRKD1*. The variant is not present in the 125,748 exomes within the GnomAD database ^20^ and has a CADD score of 29.7, thus we find it very likely that this variant is causative in our patient.

We observed enrichment of cilia genes in GS1 and we also observed enrichment of damaging RHVs in 52 cilia genes, compared to the 641 non-cilia genes in GS1. The frequency of RHVs in these 52 cilia genes are 10-fold higher in our CHD patients compared to controls. Importantly, we observed strong enrichment of cilia genes in the subset of genes with high likelihood of containing disease causing genes (GS1c) in contrast to the subset of genes with low likelihood of containing disease causing genes (GS1a), where no cilia gene enrichment was evident. We calculated the strongest enrichment in the list of 23 CDGs, wherein the frequency of cilia genes are 5.6 times higher than what would be expected by chance. Based on these results, we conclude that a significant part of disease causing genes in our patients are related to cilia structure or function. Our data thus support that cilia genes are enriched for recessive genotypes in CHD ^9^ and the high frequency of simple heart malformations in our cohort support that the involvement of cilia genes in CHD reaches beyond laterality defects, possibly due to defects in the coordination of cellular signalling by the primary cilium during development of the myocardium ^23, 24, 26^.

We identified *ADCY6* as a cilia candidate gene, with specific and strong expression in the developing heart. IFM analysis of differentiating P19CL6 cell cultures established that ADCY6 localize to the primary cilium during cardiomyogenesis and quantification of the level of ADCY6 protein within the cilium showed that the ciliary localization of ADCY6 increases at least two-fold during cardiomyocyte differentiation. The dynamic pattern of cilia localization supports an important ciliary function of ADCY6 during cardiomyogenesis. Functional investigation of zebrafish orthologues of *ADCY6* verified that the gene plays an important role in heart development.

*ADCY6* encodes an adenylyl cyclase, which have been shown to localize to the primary cilium in neuronal precursors of the developing cerebellum and neural tube, where it appears to function as an inhibitor of hedgehog (Hh) signaling ^33^. Hh signaling plays major roles in heart development and mutations of Hh signaling components have been shown to cause CHD in mouse models ^15, 34, 35^. Furthermore, recent research suggest that Hh signaling regulates the timing of differentiation of second heart field (SHF) cardiomyocyte progenitors^36^. In this context, the dynamic localization pattern of ADCY6 at the primary cilium during cardiomyocyte differentiation of P19CL6 cells might indicate that ADCY6 is involved in regulation of Hh signaling during cardiomyogenesis. However, this hypothesis needs to be addressed by further experiments using appropriate model systems for analysis of SHF progenitor differentiation.

### Strengths and Weaknesses

A strength to our work is that the genetic analysis of is based on a unique sample of patients from consanguineous families. Our analysis is based on the hypothesis that recessive inherited variants add significantly to the cause of heart malformations in patients from consanguineous families. Such recessive variants are expected to occur as homozygous genotypes in affected individuals, within HBD regions, which make up only a small fraction of an individuaĺs genome (5-6%). This reduced the number of candidate disease genes significantly, making it easier to identify potential causes. In addition, we applied a novel approach for reducing the number of candidate genes, by combining CADD score for variants with LOEUF score for genes, followed by filtering for candidate genes with specific expression in the developing heart.

The occurrence of several siblings with CHD in a consanguineous family would have supported our hypothesis, but we did only observe one affected individual in the majority of families. This could be due to various reasons, such as a possible oligogenic architecture of CHD ^7^, incomplete information regarding spontaneous abortions in the families and bias in the selection of patients, which were recruited from hospitals in major cities. Availability of WES data from the parents could have strengthened the genetic analysis by confirming HDB of variants and eliminating the possibility that some of our ROH might be regions with heterozygous genomic deletion. Pathogenic rare copy number variants (CNVs) are present in 3-10% of patients with isolated CHD ^7^, thus de novo CNVs or CNVs with penetrance in the healthy parents could in principle be causative in some of our patients. We also acknowledge the lack of an ethically matched control cohort and the use of GnomAD South Asian population as a substitute, which may not be completely representative of the Pakistani population and may not account for compound heterozygous genotypes. These limitations should be taken into consideration when interpreting our findings.

### Future Directions

Our results suggest several novel CHD candidate genes. These candidate genes need further validation in independent cohorts or by functional studies in cell or animal models.

Our data confirm a central role of the primary cilium as a signaling hub in heart development and CHD. Unravelling of the mechanisms of ciliary coordination of cell signaling in heart development remain a major challenge for understanding heart development and the pathomechanisms involved in CHD. Our current knowledge about the spatio-temporal coordination of cardiac developmental networks by the primary cilium, including regulation of crosstalk between signaling pathways, is currently very limited and experimental research addressing this field is warranted. In addition, the composition of signaling components within the cilium is very dynamic and dependent on cell-type and the surrounding tissue, which adds another level of complexity ^37^.

In summary, our genetic analysis of a cohort of CHD probands, originating from consanguineous unions, led to the identification of a 23 candidate disease genes, of which six are known ciliary genes. Our results confirm an important role for cilia genes in CHD.

**Figure 5.**
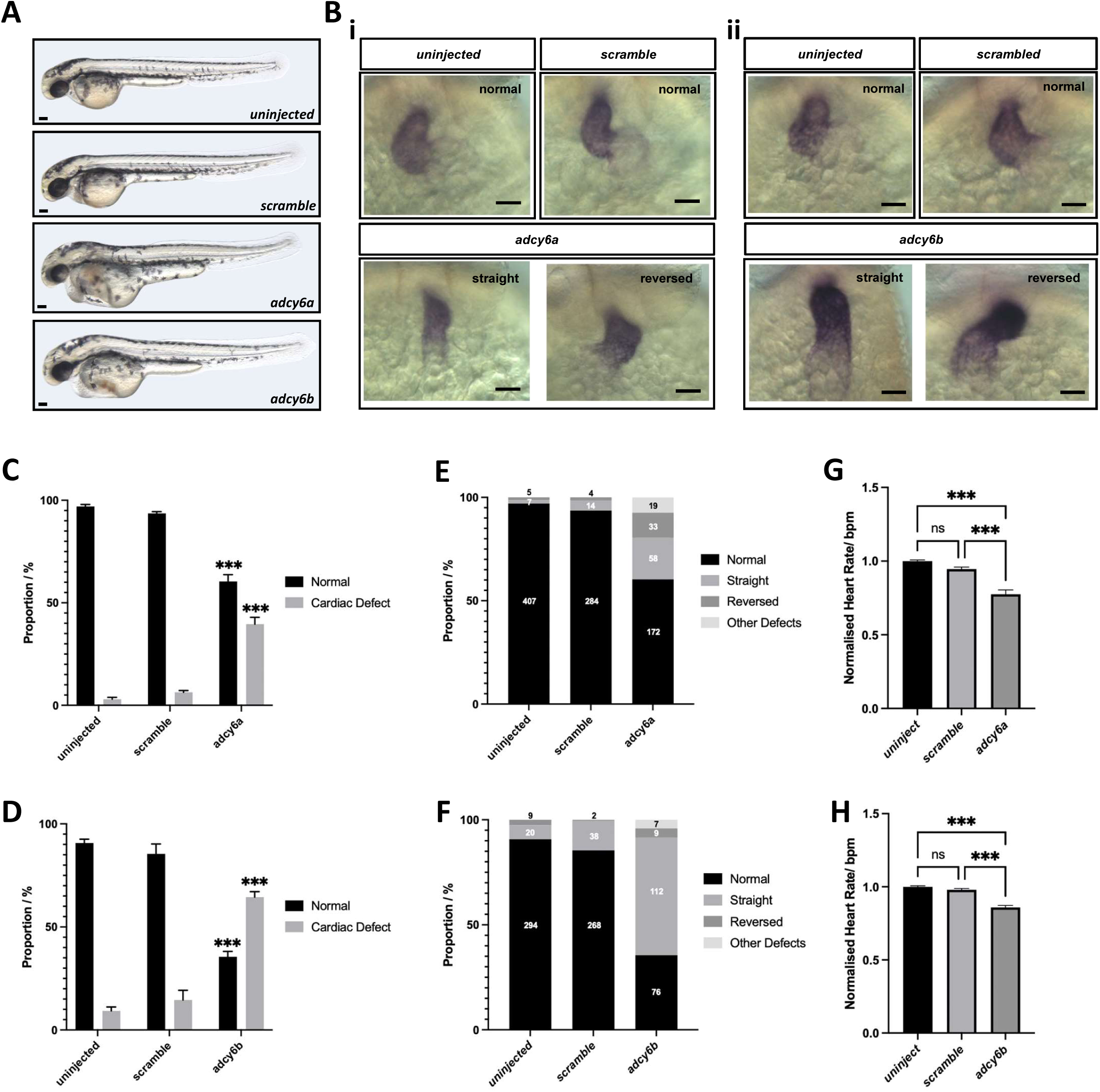
Knock-out of ADCY6 cause heart defects in zebrafish. **A**. Bright-field images showing the morphology of 2 dpf uninjected, *scramble*, *adcy6a* and *adcy6b* F0 crispant zebrafish . Scale bars, 0.5 mm. **B.** mRNA expression analysis of *myl7* in 2dpf crispant hearts. Upper panels of (i) and (ii) imaged from control larvae. Lower panels imaged from (i) *adcy6a* and (ii) *adcy6b* crispants. Scale bars, 50 µm. **C-D**. Quantification of cardiac defects observed in 2 dpf (**C)** *adcy6a* and (**D)** *adcy6b* crispants upon mRNA expression analysis of *myl7*. **E-F.** Proportion of heart phenotypes observed in **(E)** *adcy6a* and **(F)** *adcy6b* crispants. Numbers central within bars indicate number of larvae in each classification. **G-H.** Normalized heart rate measurements in beats per minute (bpm) analyzed at 2 dpf in **(G)** *adcy6a* and **(H)** *adcy6b* crispants. Two-way ANOVA (**C-D**), ordinary one-way ANOVA (**G**) and Kruskal-Wallis test (**H**) used for statistical analysis. Asterisk indicate P-values: \*\*\**p* < 0.001. n.s.: not significant.

## Data Availability

All data produced in the present study are available upon reasonable request to the authors. Individual
exome sequencing data cannot be shared due to concerns over patient privacy. Other data generated or analyzed during this study are included in the main paper or its additional files.

## Acknowledgements

We are grateful to the patients and their families for their participation in the project. We would like to thank Lillian Rasmussen for expert technical assistance.

## Supplemental Material

Supplemental Methods

Figures S1-S7

Table S1-S4

Supplemental References

