## Supplementary data for "Rare homozygous cilia gene variants identified in consanguineous congenital heart disease patients"

### SUPPLEMENTAL METHODS

#### Patients

Forty-nine patients from 48 families were recruited from cardiology departments in Rawalpindi and Lahore, Pakistan. The study followed the declaration of Helsinki and was approved by the Institutional Review Board GC University Faisalabad, Faisalabad, Pakistan. Informed consent was obtained from all participating individuals or their parents for the collection of blood samples, genetic analyses, and publication of genetic information. Patients were diagnosed by transthoracic echocardiography, cardiac catheterization, or surgery.

Consanguinity was confirmed by pedigree analysis after constructing pedigrees of each family, based on information from family elders.

#### Whole exome sequencing

Blood samples were collected from patients with an ASD and DNA was extracted using a Puregene Blood Core Kit C (Qiagen, Aarhus, Denmark). Whole exome sequencing (WES) was performed by BGI Genomics (Shenzhen, China). Coding DNA was captured using Agilent V6(60M) capture library and sequenced using PE100 sequencing on BGISEQ sequencing platforms. Sequencing-derived raw image files were processed by BGISEQ basecalling Software with default parameters and the sequence data of each individual was generated as paired-end reads and stored in FASTQ format. Clean data was produced by data filtering on raw data. All clean data of each sample were mapped to the human reference genome (hg19) using Burrows-Wheeler Aligner (BWA V0.7.15) [1]. Variant calling was performed following recommended Best Practices for variant analysis with the Genome Analysis Toolkit (GATK) [2]. Local realignment around InDels and base quality score recalibration were performed using GATK v3.7 [3, 4], with duplicate reads removed by Picard tools v2.5.0 (http://broadinstitute.github.io/picard/). All genomic variations, including SNPs and InDels were detected using GATK. Variant annotation was performed using the SnpEff tool [5]. An average of 90,345,572 clean reads (9,027.12 Mb) were obtained, with an average CG content of 54%. Mean sequencing depth of target regions was 106.1x. On average, per sample, 99.8% of targeted bases were covered by at least 1x coverage and 97.4% of the targeted bases had at least 10x coverage.

**Variant filtering**

Only homozygous variants with ≥10x coverage were included. Rare homozygous variants (RHVs) were identified according to minor allele frequency (MAF) in public databases; MAF<0.01 in 1000 genomes (total population), GnomAD total population and GnomAD south Asian population, respectively, and MAF <0.02 among the 49 patient samples. In addition, we filtered for protein altering variants (PAVs), by removing synonymous variants, intron variants (except splice region variants) and intergenic variants.

**Identification of regions of homozygosity**

Regions of homozygosity were identified from vcf files using HomozygosityMapper [6]. Setting were adjusted for identification of blocks of at least 50 homozygous variants. Circa software v.1.2.2 (OMGenomics, Redwood City, CA) was used to visualize the regions of homozygosity identified using Homozygocitymapper.

**Cilia localization of ADCY6 during cardiomyogenesis**

Temporal localization of ADCY6 to the primary cilium during *in vitro* cardiomyogenesis was evaluated by immunofluorescence microscopy analysis (IFM) in cultures of mouse embryonic P19.CL6 stem cells, which were induced to differentiate into cardiomyocytes as previously described [7]. Cells were fixed in 4% paraformaldehyde and permeabilized in 0.2% Triton X-100. Blocking was performed using 2% bovine serum albumin (BSA), and cells were incubated in primary antibodies (mouse anti-acetylated α-tubulin [1:2000, Sigma-Aldrich T6793], ADCY6 [1:500, Invitrogen PA5-118931]) overnight at 4°C. Secondary antibodies were incubated for 45 min, and 4′,6-diamidino-2-phenylindole (DAPI) staining performed prior to mounting. Cardiomyogenesis was validated by IFM using the following primary antibodies: mouse anti-SOX2 [1:400, R&D Systems, MAB2018], goat anti-GATA4 [1:200, Santa Cruz, Sc-1237], mouse anti-α-actinin [1:400, Sigma-Aldrich, A7811] and rabbit anti-Troponin-I [1:400, R&D Systems, MAB8594]. Secondary antibodies: Alexa-Fluor$^{568}$-conjugated donkey anti-mouse, Alexa-Fluor$^{568}$-conjugated donkey anti-rabbit, Alexa Fluor$^{488}$-conjugated donkey anti-mouse, Alexa-Fluor$^{488}$-conjugated donkey anti-rabbit, Alexa Fluor$^{488}$-conjugated donkey anti-goat IgG [all 1:600, Invitrogen/Life Technologies]

Fluorescence images were captured on a fully motorized Olympus BX63 upright microscope with an Olympus DP72 color, 12.8-megapixel, 4140 × 3096-resolution camera. The software used was Olympus CellSens Dimension version 1.7, which was able to do deconvolution, 3D isosurface projections on captured z stacks and slice views. Images were processed for publication using Image J version 2.0 and Adobe Photoshop CS6 version 13.0. For quantifications, the mean fluorescence values for ADCY6 at the cilium-centrosome axis were set relative to the fluorescence values in background areas of the cytosol. All data were gathered for n=3, statistical calculations were performed with the ANOVA test and data were presented as violin plots using GraphPad Prism 9 software.

**Zebrafish husbandry**

The zebrafish AB Wild-type (WT) strain was obtained from the Zebrafish International Resource Center (ZIRC). All animals were maintained in the animal facility at the University of Copenhagen, Denmark. WT zebrafish were raised in a constant light-dark cycle at 28ºC according to standard protocols. Staging and maintenance of embryos were carried out as previously described [8]. All experiments were approved and conducted according to licenses and guidelines from the Danish Animal Experiments Inspectorate (Protocol code: P20-387).

**crRNA design and selection**

crRNAs were designed for *adcy6a* (ENSDARG00000061445) and *adcy6b* (ENSDARG00000027797) and evaluated using online tools from IDT (Integrated DNA Technologies)(https://eu.idtdna.com/site/order/designtool/index/CRISPR_CUSTOM), CCTOP [9] (https://cctop.cos.uni-heidelberg.de:8043), and CRISPOR [10] (http://crispor.tefor.net). The designing of each crRNA was done according to previously described protocols [11]. Briefly, four crRNAs were designed for each *adcy6* orthologue to increase the probability of introducing biallelic frameshift mutations. The four crRNAs were designed, where possible, to target distinct asymmetrical exons whilst avoiding the first exon. Selection of crRNAs was based on the ranking of predicted on- and off-target scores. Designed guides, IDT design codes, and exons targeted for *adcy6a* and *adcy6b* are shown in Supplementary Table S3 and Figure S6A. Guides were tested for activity before experiments were conducted (Table S4 and Figure S6B).

**gRNA/Cas9 complex assembly**

Assembly of the gRNA/Cas9 complex for injection was performed as described previously [11]. In brief, equimolar volumes of crRNAs and tracrRNA were mixed in nuclease-free Duplex buffer (IDT) and incubated at 95ºC for 5 minutes to create a 57 µM crRNA:tracrRNA complex. Alt.R S.p. HiFi Cas9 Nuclease V3, 61µM (IDT) was diluted to 57 µM with Cas9 buffer (20mM Tris-HCl, 600 mM KCl, 20% glycerol). Equimolar volumes of crRNA:tracrRNA and diluted Cas9 were mixed and incubated at 37ºC for 5 minutes to formulate a 28.5µM RNP. For each *adcy6* orthologue, the four RNP solutions were mixed in equal volumes and stored at -20ºC. The same procedure was completed with three *scrambled* crRNAs (Alt-R CRISPR-Cas9 Negative Control crRNA #1, #2, #3, IDT) used as negative controls.

**Injections, genotyping and phenotyping**

Approximately 1 nL (28.5 fmol) of pooled RNP was injected into the cytoplasm of single-cell staged embryos. Survival of injected embryos were monitored several hours after injections and 1 day post fertilization (dpf). Lateral images of 2 dpf zebrafish were taken by positioning larvae with 3% methylcellulose under a Zeiss AxioZoom V16 microscope (Carl Zeiss, Brock Michelsen A/S, Denmark). Individual larvae were genotyped accordingly until it was confirmed that crispant mutants were generated in >95% of the larvae. After confirmation, larvae were genotyped in batches of 10 larvae.

**ISH in zebrafish**

Anti-sense *myl7* riboprobes were synthesized from a pGEM-T easy vector before being linearized, digoxygenin (DIG)-labeled, and transcribed with T7 RNA polymerase (Roche). ISH was performed as previously described [12] with minor modifications. Briefly, 2 dpf larvae were fixed in both 4% PFA and 100% methanol. Larvae were rehydrated in a series of dilutions and permeabilized with Proteinase K (10µg/ml) before being re-fixed in 4% PFA and pre-hybridized. Larvae were hybridized overnight at 70ºC with the *myl7* probe. Following washes, larvae were incubated overnight at 4ºC in anti-DIG antibody (1:6000, Roche) before being washed again. Probe staining was detected with NBT/BCIP (Roche) solution. Stained larvae were imaged with a Zeiss AxioZoom 16 microscope and analyzed. Larvae were subsequently genotyped as previously stated.

#### Heart rate analysis in larvae

Larvae at 2 dpf were removed from a 28ºC incubator in batches of five and allowed to acclimatize to room temperature. Larvae were anaesthetized with MS222 (1:100, Sigma-Aldrich) for 3 minutes. Positioning of the larvae in a brightfield microscope was done so that the heart was clearly visible. The heart rate for 15 seconds was then manually counted. Heart rates for one minute were subsequently calculated and larvae were genotyped as previously stated.

#### Statistical analyses

Statistical analyses were performed using Graphpad Prism v.9.5. For data in Fig 3E and Supplementary Figure S3, a Mann-Whitney U test was performed to obtain exact P-values (https://www.statskingdom.com/170median_mann_whitney.html). Adjustment for multiple testing was performed with the Benjamini-Hochberg method (https://tools.carbocation.com/FDR). Enrichment of known CHD genes was determined by calculating overlap between gene lists. Significant overlap was calculated using hypergeometric statistics (http://nemates.org/MA/progs/overlap_stats.html). A representation factor was calculated as the number of overlaping genes, divided by the expected number of overlapping genes drawn from two independent groups; RF=x/((n*D)/N), where x=number of overlapping genes, n=genes in group 1, D=genes in group 2, N=genes in genome (20,000).

Data from zebrafish experiments are presented as the mean ± SEM. All results from the experiments were validated by independent repetitions and were carried out with n ≥ 3. Representative images and graphs are presented. Data was tested for Gaussian distribution with a Shapiro-Wilk test. Statistical difference between groups was completed by either ordinary one-way analysis of variance, ordinary two-way analysis of variance or with a Kruskal-Wallis test.

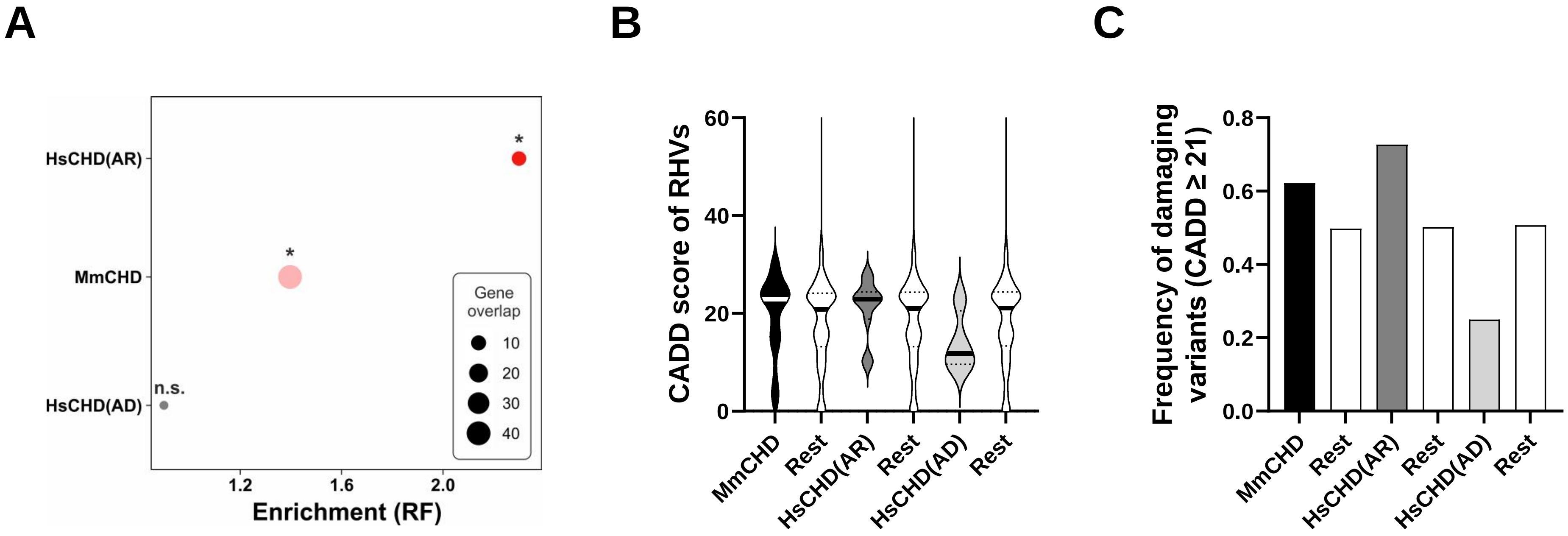


**Figure S1. Enrichment of known CHD genes among 693 genes with RHVs. A.** The overlap between lists of genes known to cause CHD in humans and mice and the list of 693 genes in GS1 (Table S1) is shown as circles. Enrichment was calculated using a hypergeometric test. Enrichment is shown as representation factor (RF). Statistical significance of the overlap is indicated. *: P<0.05, ns: not significant. HsCHD(AR) : a list of genes known to cause autosomal recessive CHD in humans (N=115)[18], MmCHD(AR): a list of genes known to cause CHD in recessive mouse models (N=832)[17], HsCHD(AD): a list of genes known to cause autosomal dominant CHD in humans (N=130)[18]. B, C. Distribution of CADD scores of gene variants (B) and frequency of damaging variants (C) in overlapping genes in A, compared to the GS1 genes that are not overlapping (Rest). An ANOVA test determined that there is no statistical difference between medians (B) or frequencies (C).

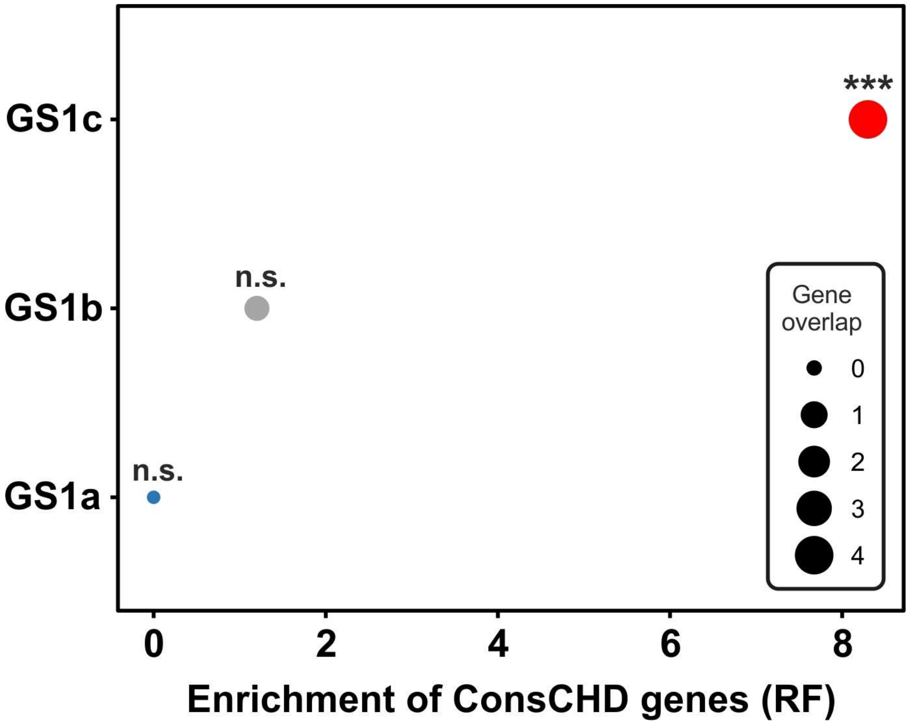


**Figure S2. Enrichment of human CHD gene-set obtained from clinical exome sequencing of CHD patients from a consanguineous cohort.** The set of 50 genes (ConsCHD) is listed in Supplementary Table S2. Enrichment was calculated by comparing gene-overlap between ConsCHD genes and the three sub-groups of CDGs. Enrichment is shown as representation factor (RF). A hypergeometric distribution was used to test the significance of the overlaps. Asterisks indicate P-values: *** P<0.001, n.s.: not significant.

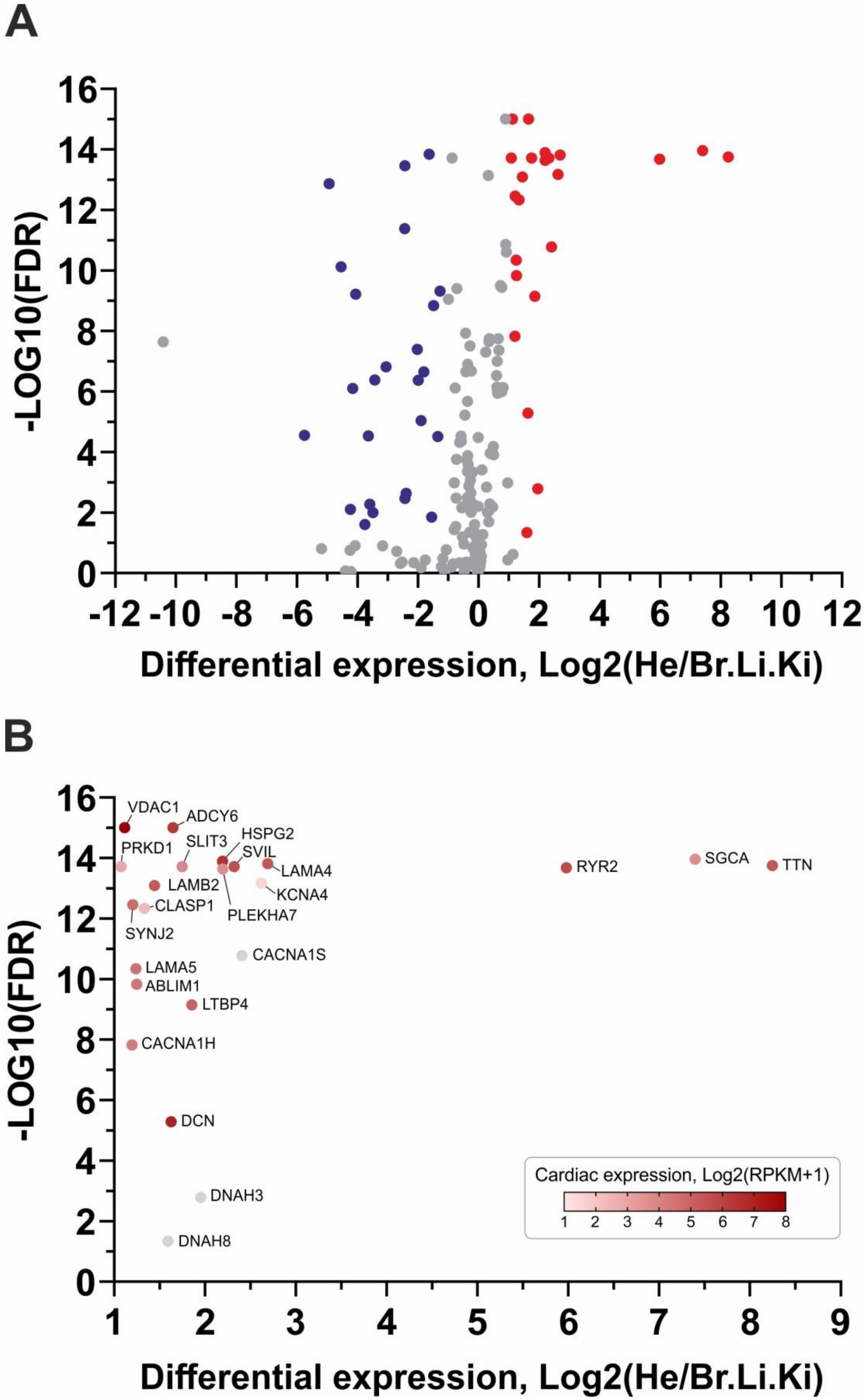


**Figure S3. Cardiac specific expression of GS1c geneset. A.** Vulcano plot of genes in GS1c geneset showing the differene in gene expression between developing heart (He) and developing brain (Br), Liver (Li) and Kidney (Ki) in mice at E10.5-E18.5 . X axis shows the log2 difference between average expression in He and average expression in Br, Li and Ki. Y axis shows the significance, calculated as –Log10 to the false discovery rate (FDR) (Mann-Whitney U test, adjusted for multiple testing). Significant genes, with fold change >1 and <-1 is shown with red and blue color, respectively. **B.** Vulcano plot of 23 significant genes in (**A**) with fold change >1. The color of the circle indicate Log2 of the average expression of the gene in developing hearts. Expression data was obtained from [22].

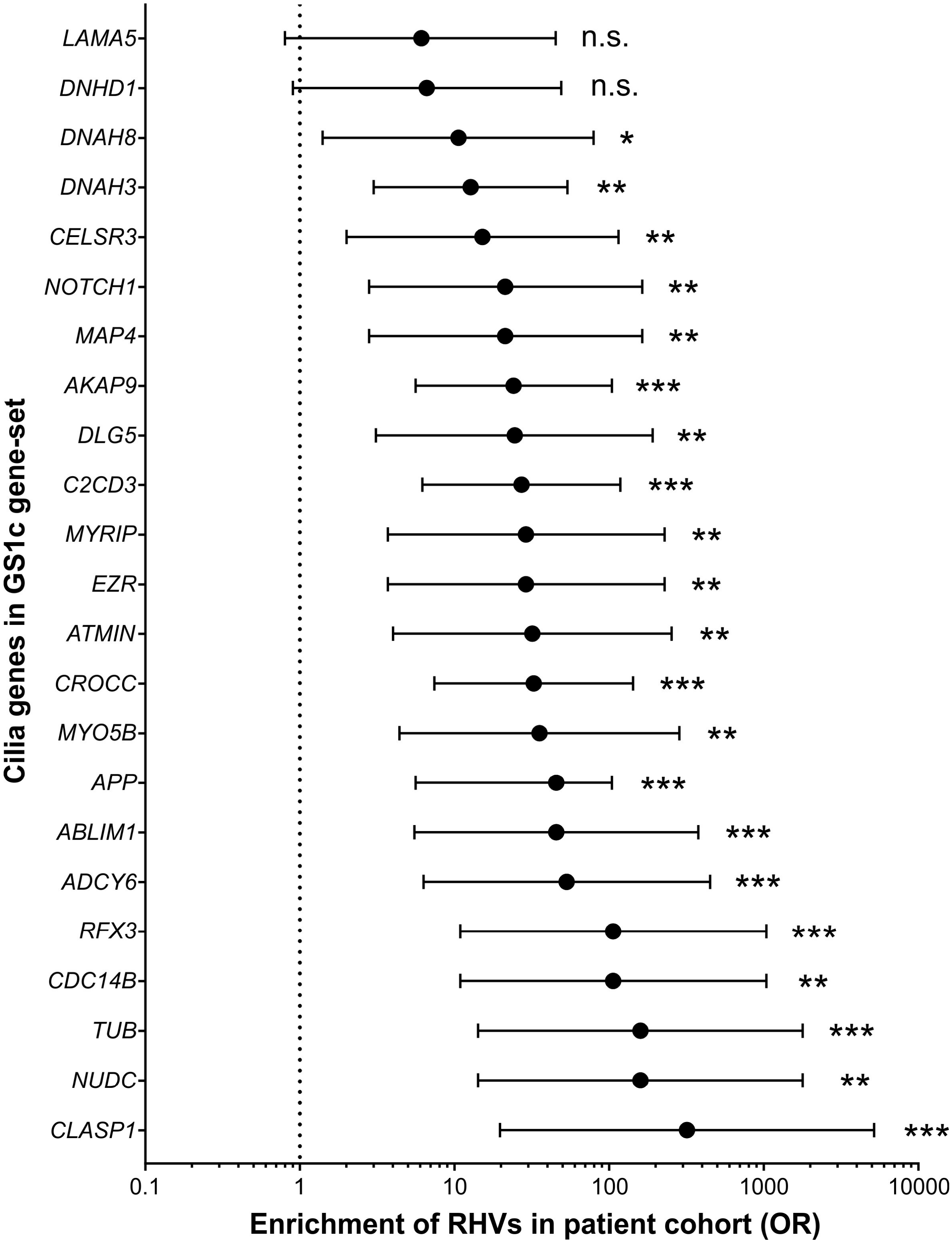


**Figure S4. Forrest plot of Enrichment of RHVs in each of 23 cilia genes in our patient cohort.** The frequency of RHVs in patients were compared with the frequency in 15,308 South Asian controls (GnomAD v2.1.1). Enrichment was calculated as an odds ratio (circles)(medcalc.org/calc/odds_ratio). The 95% confidence interval is shown with bars. Asterisks indicate P-values: * P<0.05, ** P<0.01, *** P<0.001. n.s.: not significant.

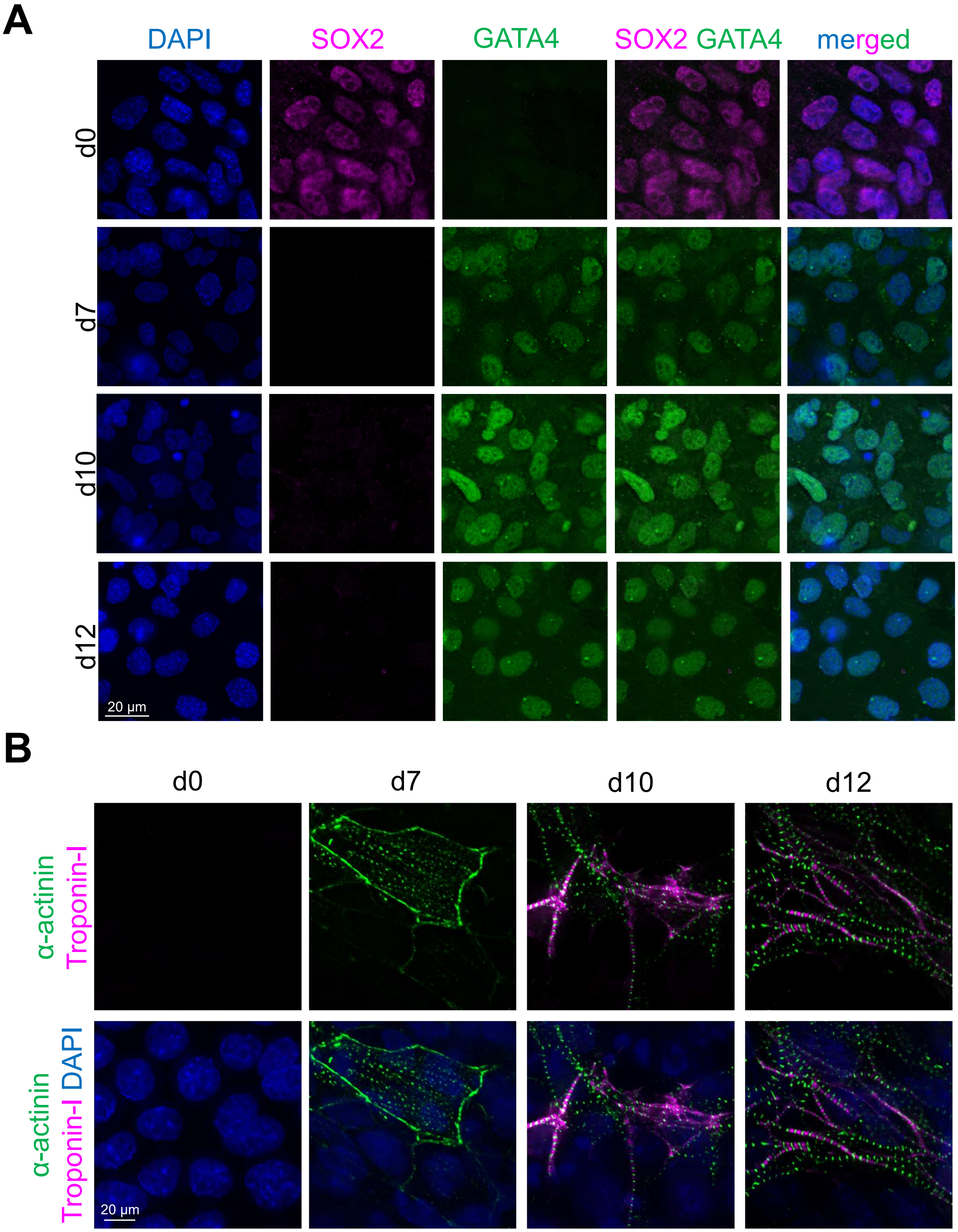


**Figure S5. Cardiomyogenesis of P19CL6 cells.** Immunofluorescence microscopy (IFM) analysis of cells after addition of DMSO. Samples were analysed at day 0 (d0), day 4 (d4), day 7 (d7), day 10 (d10) and day 12 (d12). A. IFM using antibodies against stem cell marker SOX2 (magenta) and the cardiac transcription factor GATA4 (green). **B.** IFM using antibodies against Troponin-I (magenta) and α-actinin (green) Nuclei were stained with DAPI (blue).

A

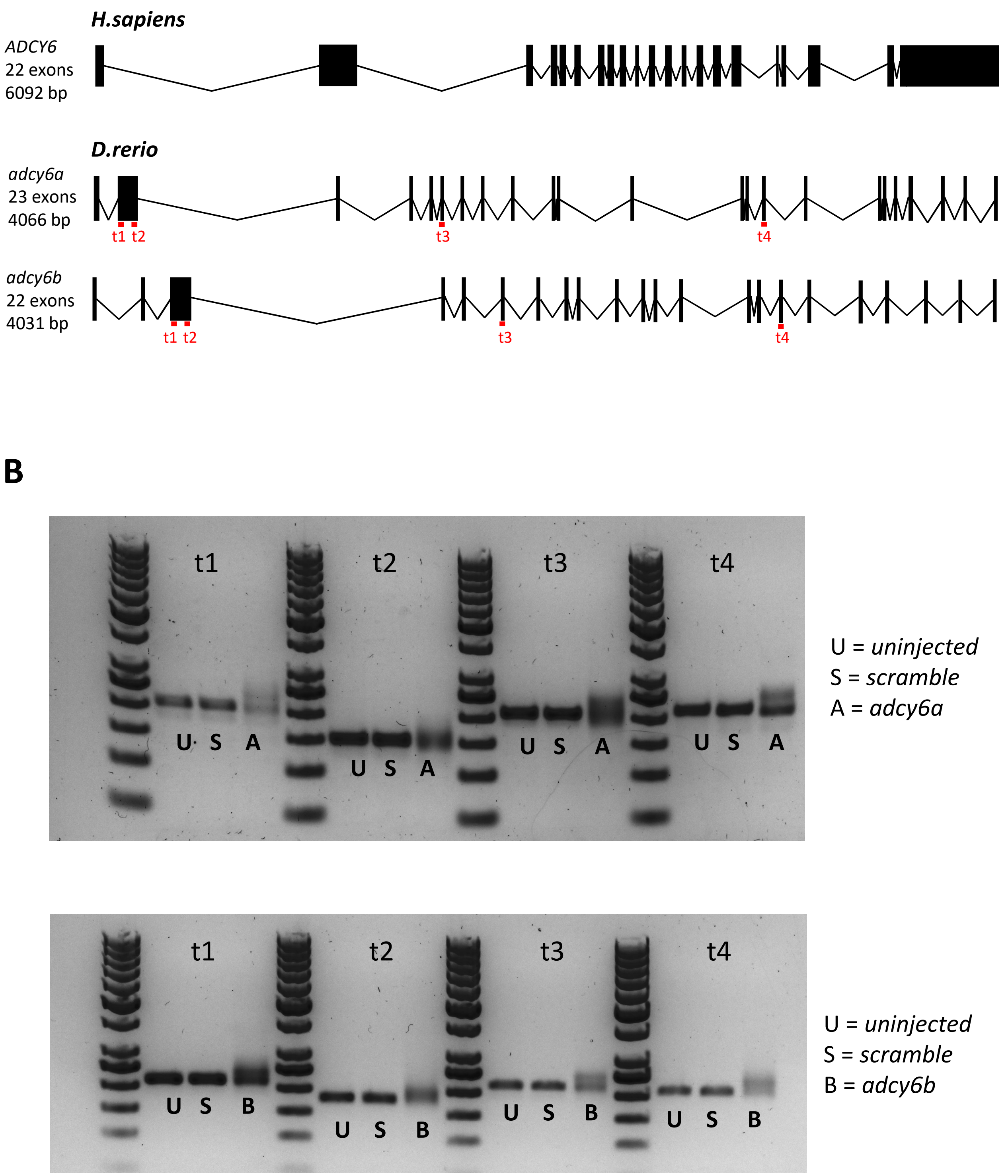


**Figure S6. CRISPR-cas9 mediated mutagenesis of *adcy6* in F0 zebrafish embryos (crispants). A.** Four guides were designed to target each of the zebrafish *adcy6a* and *adcy6b* gene, respectively. The zebrafish ortologues are shown below the human *ADCY6* gene. Exon sequence is indicated with black squares, intron sequence is indicated with lines. Target sites (t1-t4) of guide RNAs are shown with red lines. **B.** Example of genotyping gel for analysis of mutagenesis efficiency in crispants.

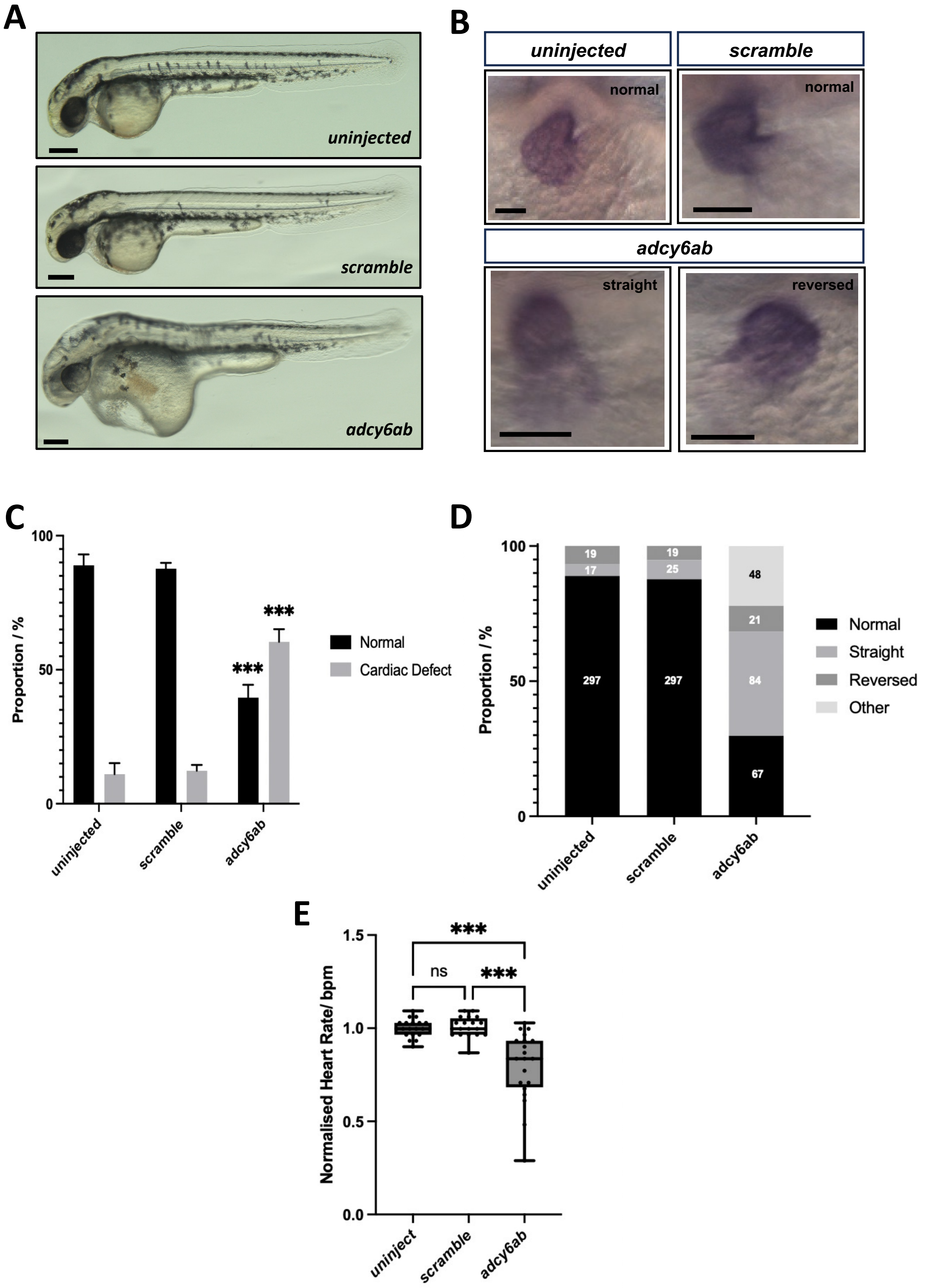


**Figure S7. Double knockout of *adcy6a* and *adcy6b* causes heart defects A**. Bright-field images showing the morphology of 2 dpf uninjected, scramble, and *adcy6ab* F0 crispant zebrafish larvae. Scale bars, 0.5 mm. **B**. mRNA expression of *myl7* in 2 dpf crispant hearts. Upper panels show control larvae. Bottom panels show *adcy6ab* crispants. Scale bars, 100 µm. C. Proportions of cardiac defects observed in 2 dpf mRNA expression analysis of *myl7*. **D**. Proportion of cardiac phenotypes observed in 2 dpf F0 crispants. Numbers central within bars indicate number of larvae in each classification. **E**. Normalized heart rate measurements for 2 dpf F0 crispants in beats per minute (bpm). Two-way ANOVA (**C**) and ordinary one-way ANOVA (**E**) used for statistical analysis. Asterisk indicate P-values: *** P<0.001, n.s: not significant.
